# Age affects temporal response, but not durability, to serial ketamine infusions for treatment refractory depression

**DOI:** 10.1101/2020.08.31.20185538

**Authors:** Steven Pennybaker, Brian J Roach, Susanna L Fryer, Anusha Badathala, Art W Wallace, Daniel H Mathalon, Tobias F Marton

**Affiliations:** Department of Psychiatry and Behavioral Sciences, University of California, San Francisco; San Francisco VA Health Care System; Department of Anesthesiology, University of California, San Francisco

**Author notes:** Drs. Fryer, Wallace, Mathalon and Marton are U.S. Government employees. The content is solely the responsibility of the authors and does not necessarily represent the views of the Department of Veterans Affairs. Corresponding author: Tobias F. Marton, MD, PhD, University of California, San Francisco, Department of Psychiatry and Behavioral Sciences, Weill Institute for Neurosciences, San Francisco VA Health Care System/Mental Health Service (116D), 4150 Clement Street, San Francisco, CA 94121.

**Keywords:** major depressive disorder, treatment-resistant depression, ketamine, antidepressant, pharmacology

## Abstract

**Background:** Ketamine is a rapid-acting treatment for patients with treatment refractory depression (TRD), however treatment responses are often transient and ketamine’s antidepressant action lacks robust clinical durability. Little is known about which patient characteristics are associated with faster or more durable ketamine responses. Ketamine’s antidepressant mechanism is proposed to involve modulation of glutamatergic signaling leading to long term potentiation (LTP) and synaptogenesis, and these neuroplasticity pathways have been shown to be attenuated with older age. We therefore investigated the impact of patient age on the speed and durability of ketamine’s antidepressant effects in veterans receiving serial intravenous ketamine infusions for TRD.

**Methods:** Beck Depression Inventory (BDI-II) scores from 49 veterans receiving six ketamine infusions (twice weekly) were examined from a retrospective case series. Percent change in BDI-II scores across the infusion series were assessed with respect to patient age using a mixed-linear model. Follow-up analyses examined the age x infusion number interaction effect at each assessment time point. To assess treatment durability, BDI-II change scores three weeks following the sixth infusion were correlated with age.

**Results:** There was a significant age x infusion number interaction (*F*=3.01, *p*=.0274) across the six infusions. Beta estimates at each infusion showed a significant effect of age at infusion #4 (*B*=.88% +/-.29%, *t*=3.02, *p*=. 004) and a trend towards significance at infusion #5 (*B*=.62% +/-.31%, *t*=1.95, *p*=.057). There was no significant correlation between percent change in BDI-II and age at three-week follow-up.

**Conclusions:** Older age is associated with an altered trajectory of antidepressant response across serial ketamine infusions, with a model-predicted difference of 8.8% less improvement in BDI-II score for each decade in age mid-way through the infusion course. In contrast, antidepressant durability at three-week follow-up was not related to age. These data suggest age is an important moderating factor of patient response to ketamine, and that differing mechanisms may underlie speed and durability of ketamine’s antidepressant activity.

## Introduction

Treatment refractory depression (TRD), commonly defined as failure to respond to at least 2 antidepressant trials of adequate dose and duration, effects up to one third of patients with Major Depressive Disorder (MDD) and is associated with greater rates of relapse, prolonged disability, higher medical costs, and lower life quality then treatment-responsive MDD^1-3^.

The dissociative anesthetic ketamine has garnered increasing enthusiasm as a highly effective and rapidly acting antidepressant with a novel mechanism of action^4,5^. A single sub-anesthetic infusion of ketamine results in remission from depression in 40 to 80% of TRD patients within 3 to 72 hours of treatment^6,7^. However, these impressive antidepressant effects are ephemeral with a majority of patients fully relapsing within one week of treatment^6,8^. While providing multiple infusions in series can prolong treatment efficacy to an extent^9,10^, antidepressant durability remains a significant limitation of ketamine therapy as a treatment modality. Further, the parameter space of ketamine delivery for MDD (i.e. optimizing dosing, number of treatments, treatment interval etc.) has been largely unexplored^5^, with most multi-infusion studies examining the effects six serial ketamine infusions^9-11^. Among other important directions, a more complete understanding of the individual factors that moderate treatment response is a critical step towards improving, optimizing and ultimately personalizing ketamine therapy for TRD.

Ketamine’s rapid and highly effective antidepressant action is unique amongst existing antidepressant therapies. While ketamine’s precise mechanism of action is unclear, current research supports the likely importance of its action as an inhibitor of the N-methyl-D-Aspartate receptor (NMDAR)^12-15^, which is necessary for glutamatergic signaling, long-term potentiation (LTP) and neuroplasticity^16^. Impairments in LTP and NMDAR functioning are increasingly recognized as a potential common pathophysiological mechanism for numerous psychiatric disorders including schizophrenia, anxiety disorders, addiction, and mood disorders, including MDD^16,17^.

Numerous lines of preclinical and clinical evidence support impaired glutamatergic signaling, NMDAR functioning and LTP in MDD^15^. This glutamatergic view has challenged the classical “monoamine hypothesis” of MDD, in which reductions in monoamine neurotransmitter levels (e.g. serotonin, dopamine) are central to the pathophysiology of depression (and corrected by SSRI administration)^17^; Instead, derangements in glutamate signaling leading to attenuated LTP-based neuroplasticity in mood-regulation networks may represent a more fundamental, downstream pathophysiological mechanism, and therefore a more direct target for antidepressant therapeutics such as ketamine^4,15^.

Age has been shown to be a moderating factor of response to numerous antidepressant treatments, including SSRIs and intranasal esketamine^18,19^. Further, advancing age has also been shown to attenuate LTP and related neural plasticity mechanisms in preclinical models^20^, and these findings have been extended to humans, showing reduced cortical plasticity as measured by both motor evoked potentials^20,21^ and EEG-based measures^22,23^.Ketamine also activates and upregulates α-amino-3-hydroxy-5-methyl-4-isoxazolepropionic acid (AMPA) receptors, as well as synaptogenic signaling pathways^24^. This is notable, given that AMPA receptor downregulation and decreased synaptogenesis is associated with aging^25^.

Given the strong implication of LTP-like and associated glutamatergic signaling underlying ketatmine’s antidepressant mechanism, as well as evidence of age-related decline in neuroplasticity, we examined relationships between patient age and treatment response trajectories and response durability in a case-series retrospective analysis of 49 veterans with TRD treated with a series of six ketamine infusions at a VA medical center. Prior studies examining the impact of age on ketamine’s antidepressant efficacy represent a modest literature mostly consisting of small case series and case reports with some conflicting results. Several, but not all^26^,case series examining ketamine’s efficacy in older patients report antidepressant efficacy similar to non-geriatric patients^27,28^, and a small randomized-controlled trial of subcutaneous ketamine also demonstrated significant effects of ketamine in geriatric patients^29^. However, the Transform-3 trial, a large multi-site RCT that assessed the efficacy of intranasal esketamine in patients over 65 with TRD, failed to show separation from placebo following 4 weeks of twice-weekly treatment, though *post-hoc* analysis of patients between 65 and 74 did demonstrate efficacy at this time-point, suggesting possible effects of age on esketamine treatment response and/or response trajectory^18^.

Together, these data suggest that age may be an important factor to consider when using ketamine to treat TRD in older patients, but that additional studies are needed to fully explore this important issue. We therefore examined this question in our retrospective sample and hypothesized that age would be a moderating factor of treatment trajectory in veterans with TRD receiving a course of six serial ketamine infusions.

## Methods

### Participants and Procedures

We examined retrospective treatment outcome data of 49 consecutive veterans treated with intravenous ketamine for TRD at the San Francisco VA Medical Center. Inclusion criteria included two or more failed antidepressant trials, moderate-to-severe current depression severity, and ability to give informed consent to the clinical treatment; exclusion criteria included DSM-5 Psychotic Disorders or current substance use disorder, other than cannabis. All patients received six infusions of ketamine (0.5 mg/kg over 40 minutes) twice weekly for 3 weeks in the post-operative care unit (PACU) of the San Francisco VA Medical Center. The majority of patients received the 0.5 mg/kg dose for all 6 infusions. However, seven patients had their ketamine dose increased to 0.6 mg/kg (n=6) or 0.7 mg/kg (n=1) over the six-infusion course. Ketamine infusions were provided under the care of both an anesthesiologist and psychiatrist and with standard vital sign monitoring (heart rate, blood pressure and oxygen saturation monitoring). Beck Depression Inventory-II (BDI-II) data were collected immediately prior to each infusion.

Following the initial induction phase of six infusions, 36 of the 49 patients progressed to a maintenance phase of treatment consisting of one ketamine infusion every 3 weeks. To investigate the antidepressant durability of the initial induction phase of six infusions, we also examined follow-up BDI-II data collected directly prior to the first maintenance ketamine infusion, which was 3 weeks following the final (sixth) of the initial infusion series (3-week follow-up).

### Statistical Analysis

Mixed Linear Model: To test our central hypothesis that trajectories of clinical response vary as a function of patient age, we employed a mixed model in SAS (v9.4), with BDI-II percentage change from baseline as the dependent variable, Time (infusions #2-6) as a repeated measure, Age as continuous covariate, and subject as a random factor. We considered compound symmetric, auto-regressive (AR-1), and unstructured covariance structures to account for the correlated nature of the BDI-II percent change repeated measure, with the final model selected based on best Bayesian Information Criteria (BIC)^30^. We first examined the main effect of Time (with Tukey-Kramer adjusted follow-up comparisons), in order to characterize the clinical response trajectory of the entire cohort, over the infusion series. Next, our main effect of interest, the Age X Time interaction effect was parsed with planned follow-up tests assessing the relationship between BDI-II percent change and age at each time point. For all follow-up tests, type 1 error was controlled at p<.05, familywise.

Treatment Durability: Durability of response to ketamine was assessed with a paired t-test examining the difference in BDI-II percent change from Infusion #6 versus the change at 3-week follow-up (both relative to pre-infusion #1 BDI-II). Lastly, to further examine age effects in our data, age was correlated with durability of treatment response.

## Results

### Demographic and Clinical Characteristics

The mean age of the patient cohort was 52.5 with a range from 22 to 77 years old. 45 of the 49 patients were diagnosed with treatment-resistant MDD and four patients with bipolar disorder, currently in a depressive episode. The patients were largely male (75%) and had a high rate (34%) of comorbid post-traumatic stress disorder (PTSD), consistent with patient demographics of the veteran population. The sample also had a high rate of treatment resistance, with 9.2 mean failed med trials and 36.7% having previously failed ECT. Patient demographic information is summarized in **Table 1**.

**Table 1.** Demographic and Clinical Data.

| Clinical Data | Patients |
| --- | --- |
| *n* | 49 |
| Age (years; range 22 to 77) | 52.5±14.2 (SD) |
| Gender (% Male) | 75% (37/49) |
| # of prior antidepressant medication trials | 9.2 ±4.4 |
| Prior ECT (%) | 36 (18/49) |
| Comorbid PTSD (%) | 34 (17/49) |
| Bipolar, diagnosis (%) | 8.2 (4/49) |
| BDI-II score pre-ketamine | 35.5±11.2 |
| BDI-II score at infusion #6 | 16.8±12.3 |

Abbreviations: BDI-II, Beck Depression Inventory version 2. ECT, electroconvulsive therapy. PTSD, post-traumatic stress disorder. SD, Standard Deviation.

### Age Effects on Clinical Response Trajectory and Durability

For the final linear mixed model, an unstructured covariance matrix was selected, based on BIC^30^. The model revealed a significant Age X Time interaction effect (F(4,47)=3.01, *p*=0.027), a main effect of Time (F(4,47)=3.49, *p*=0.0142) but no main effect of Age (F(1,47)=3.04, *p*=0.0876). We first examined the main effect of time to establish the pattern of clinical treatment response across the entire sample, collapsed across age. The BDI-II assessed just prior to infusion #1 was used as the reference to calculate percent change scores with each subsequent infusion (i.e. percent change score for infusion #2 would represent the change score from infusion #2 to #1, thus reflecting the effects of a single infusion, and so on for the six infusion series). The main effect of Time across the infusion series was parsed with Tukey-Kramer adjusted follow-up comparisons. **Figure 1**. shows key comparisons of these BDI-II percent change scores, including significantly greater reduction at infusion #2 compared to infusion #1, and infusion #6 compared to infusion #1, reflecting significant clinical response after both the first and final infusion (corrected p<.001).

**Figure 1.**
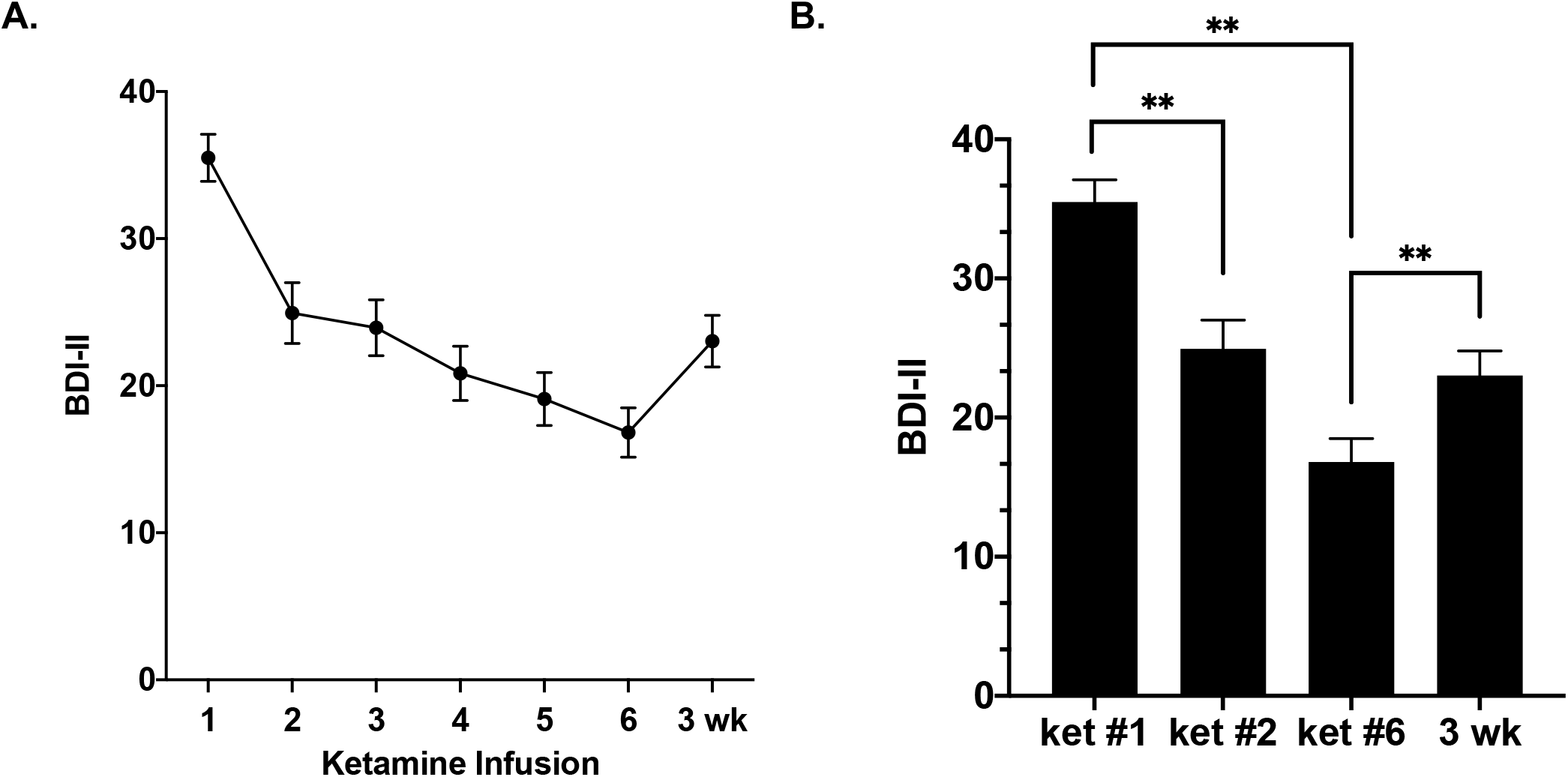
Cohort-level clinical response trajectory to the series of six ketamine infusions. Mean BDI-II scores collected prior to each ketamine infusion (#1- #6) and at 3-week follow-up (3 wk). B. Bar graphs of mean BDI-II scores collected just prior to ketamine infusion #1 (ket #1), infusion #2 (ket #2), infusion #6 (ket #6), and at 3-week follow-up (3 wk). BDI-II scores: Ket #1: 35.5+/-1.6, Ket #2: 24.9+/- 2.09, Ket #6:16.8+/-1.6, 3 wk: 23+/-1.7. ** *p < 0.001*.

Next we examined the Age x Time interaction effect. Planned follow-up tests parsed the significant interaction effect by assessing the BDI-II percent change relationship with age at each time point. There was a significant effect of age on BDI-II percent change pre-infusion #4 (*B*=.88% +/-.29%, *t*=3.02, *p*=. 004) and a trend towards significance pre-infusion #5 (*B*=.62% +/-.31%, *t*=1.95, *p*=.057) with older patients showing less percent change (i.e., older age is associated with less reduction in depression severity at infusion #4). These relationships are summarized in **Table 2**. For example, a decade increase in age corresponds to a model predicted 8.8% less percent reduction in BDI-II score pre-infusion #4. Notably, there was no significant effect of age at infusion #6 (*B*=.37% +/-.3%, *t*=1.23, *p*=. 225) indicating that patient age did not affect the total antidepressant response to the infusion series. Example modeled estimates of BDI-II response trajectories by various ages representative of the sample are shown in **Figure 2**.

**Table 2.** Model estimates of effects of age on BDI-II percent change at each infusion.

| Estimates |  |  |  |  |  |  |  |  |
| --- | --- | --- | --- | --- | --- | --- | --- | --- |
| Label | Estimate | SE | DF | t Value | Pr > \|t\| | Alpha | Lower | Upper |
| 1 year change in age at infusion 2 | 0.2225% | 0.3403% | 47 | 0.65 | 0.5163 | 0.05 | -0.4620% | 0.9071% |
| 1 year change in age at infusion 3 | 0.2207% | 0.5923% | 47 | 0.37 | 0.7111 | 0.05 | -0.9710% | 1.4120% |
| 1 year change in age at infusion 4 | 0.8861% | 0.2929% | 47 | 3.02 | 0.004 | 0.05 | 0.2968% | 1.4750% |
| 1 year change in age at infusion 5 | 0.6202% | 0.3176% | 47 | 1.95 | 0.0568 | 0.05 | -0.0190% | 1.2590% |
| 1 year change in age at infusion 6 | 0.3745% | 0.3049% | 47 | 1.23 | 0.2254 | 0.05 | -0.2390% | 0.9878% |

**Figure 2.**
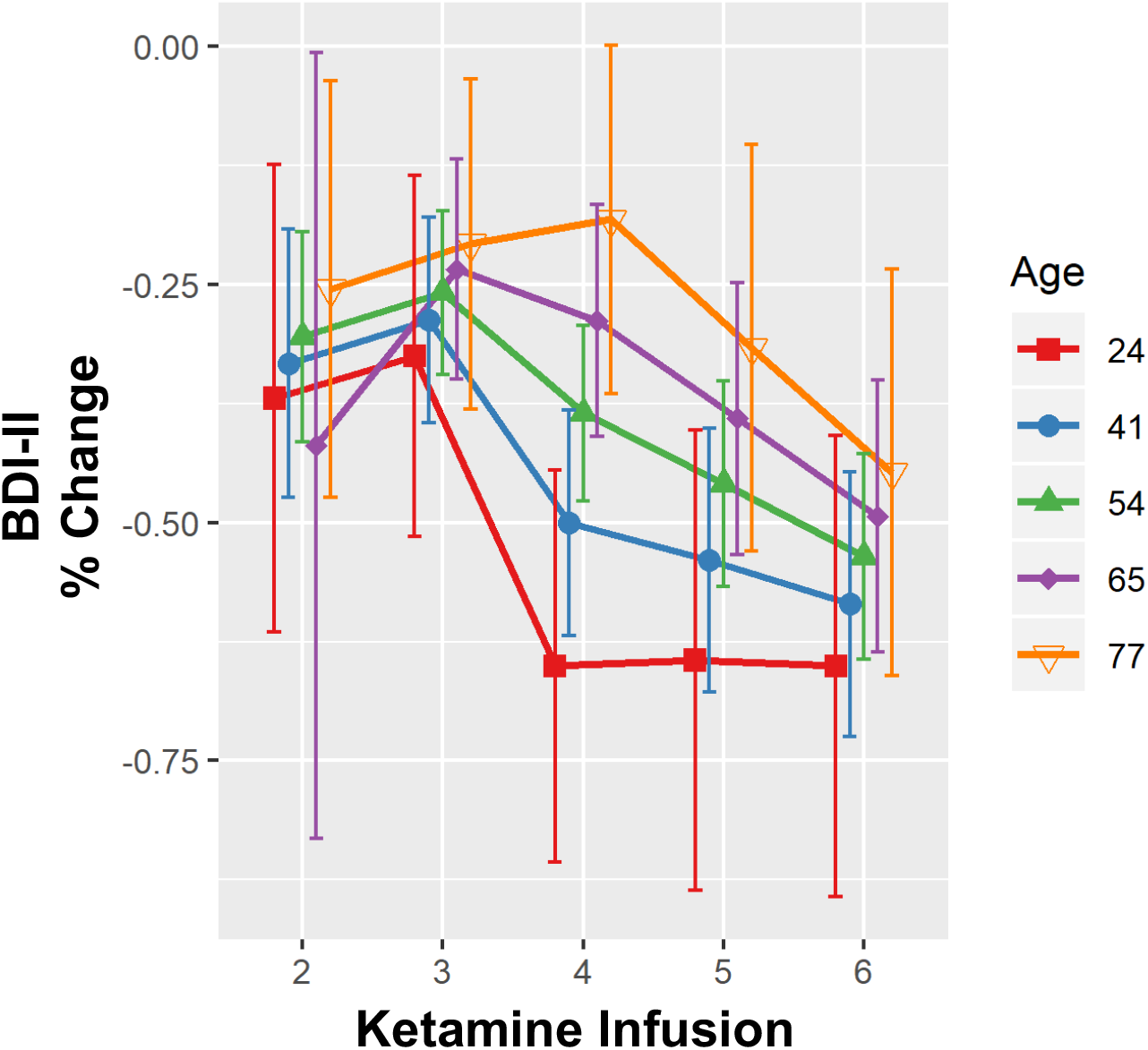
Model-Estimated trajectories of response at various ages over the infusion series. Sample response trajectories (% change in Beck Depression Inventory, BDI-II), as estimated by the linear mixed model, over the ketamine infusion series at select ages representing the minimum (22), maximum (77), and quartiles (41, 54, 65) of the patient cohort. See Table 2 for statistical analysis.

In contrast to the significant age interaction effect observed across the six-infusion series, there was no significant correlation between change in BDI-II measured at ketamine infusion #6 to the BDI-II measurement at three-week follow-up. (r (36) = 0.0005, p = 0.997) see **Figure 3**. The lack of age relationship in treatment durability was observed in the context of a significant increase in BDI-II percent change, t(35) = 3.55, p = 0.0011 (reflecting a partial relapse in depression severity from the Infusion #6 to the 3-week-follow-up, see figure 1b). These data show that while at the cohort level there was a clinically relevant change in BDI-II from infusion #6 to the 3-week follow-up assessment, age had no effect on antidepressant durability measured 3 weeks following the completion of the infusion series.

**Figure 3.**
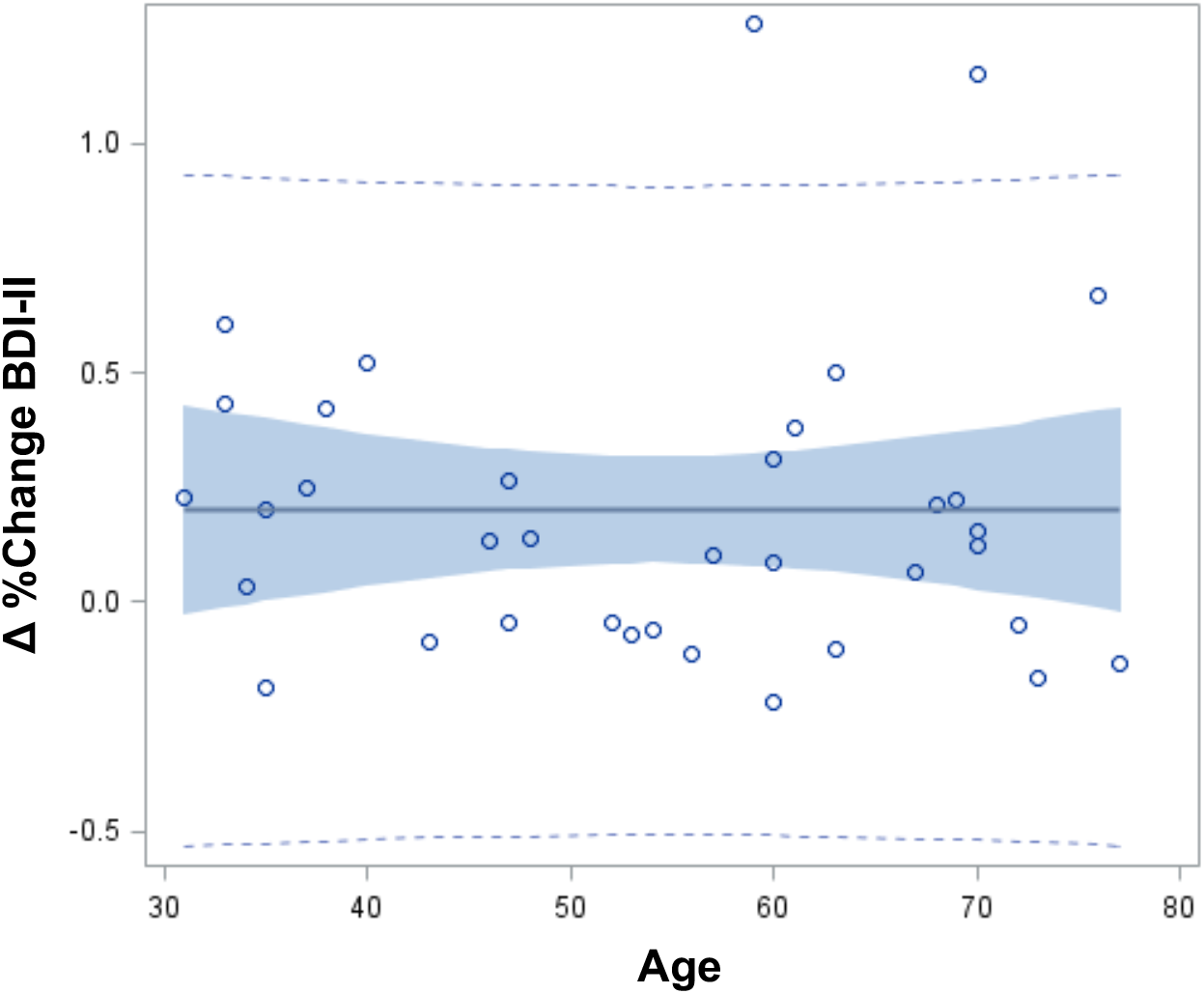
Age effects on durability of antidepressant response. Scatterplot of age x Δ percent change in Beck Depression Inventory (BDI-II) from infusion #6 to 3-week follow-up (percent change at infusion # 6 from baseline – percent change at 3 weeks from baseline) representing the durability of the antidepressant response at 3 weeks following the completion of the infusion series. Blue shading: 95% confidence interval. Correlation n.s. *n* = 36.

Finally, as seven patients had their ketamine dose adjusted during the infusion series (see Methods), we re-ran the model with these seven patients excluded to confirm our reported effects were not confounded by differences in ketamine dose and found an almost identical pattern of statistical effects with the one difference being that the BDI-II percent change – age relationship pre-infusion #5 strengthened from a statistical trend in the whole sample to reach statistical significance in the sample restricted to patients that received 0.5mg/kg dosing.

## Discussion

This retrospective analysis of the clinical response of 49 veterans receiving six serial ketamine infusions shows that older age moderated ketamine’s antidepressant effects at infusion #4 (with a trend towards significance at infusion #5), but had no effect on the efficacy of infusions earlier or later in the treatment course, nor on antidepressant durability at a three week follow-up assessment. In sum, these results suggest that age affects the trajectory of antidepressant response to serial ketamine infusions, such that older patients may respond later in the treatment course, but ultimately experience the same degree of improvement as younger patients following the completion of the six-infusion series. Elucidating this delayed-efficacy phenomenon in older patients receiving serial ketamine infusions for TRD has important implications for personalizing treatment as well as counseling patients appropriately concerning how quickly they can expect to receive benefit over the course of a multi-infusion ketamine treatment.

Further, these data reveal several interesting observations that could have implications concerning ketamine’s mechanisms of action that may inform future mechanistic studies. Based on the trajectories of response described in this study, we propose three distinct response features to an index treatment of six serial ketamine infusions: early infusion responses, late infusion responses and durability of response at 3-week follow-up. Further, we propose that the clinical phenomenology described here suggests that these three phases of response to ketamine may have different mechanisms.

Age had no effect on the clinical response to infusions early in the treatment series. It is of interest to note that ~30% of the total group clinical response to the six-infusion series occurred following ketamine infusion #1 (with the remaining response evolving more slowly over the subsequent infusions) (see **Figure 1a,b**). This is both broadly consistent with the extensive literature examining the efficacy of a single ketamine infusion, and with existing literature showing antidepressant response trajectories to a six-infusion series^9-11^

The fact that the first ketamine infusion drives ~30% of the total clinical response (i.e., percent change in BDI-II scores) to the infusion series, and that age only moderates antidepressant efficacy to infusions later in the treatment course, suggests that distinct mechanisms may drive effects of early versus later ketamine infusions in the treatment series. For example, as existing lines of evidence demonstrate reductions in neural plasticity with advancing age^20,23,25^, one possible interpretation of our data is that LTP and glutamatergic mechanisms are more important later in the infusion course as compared to earlier infusions. What mechanisms would then be predominant for earlier infusions? Recent work by Williams et al.^31^ showed that pre-treatment with opioid receptor antagonist naltrexone resulted in attenuated efficacy of a single ketamine infusion, arguing that ketamine’s antidepressant mechanisms may be largely mediated via the opioidergic system. Interestingly, other published case series data, including work from our group, showed no effect of concurrent opioid receptor modulator use or naltrexone on the response to serial ketamine infusions^32,33^. Perhaps ketamine’s antidepressant mechanism is more opioid-dependent early in treatment, and plasticity mechanisms contribute more with successive infusions (or as a function of time). Our results contribute to a body of work that suggests a possible framework with which to design mechanistic studies to parse the relative contributions of neural plasticity and opioidergic signaling in ketamine’s antidepressant action.

Our finding that age has no effect on the durability of ketamine’s antidepressant response (at 3 weeks) is also informed by recent literature and raises further mechanistic questions. D-cycloserine is a partial agonist at the glycine site of the NMDAR and has been shown to have pro-plasticity effects in preclinical models and in clinical studies^34,35^. However, d-cycloserine administration following a single ketamine infusion failed to extend antidepressant durability in a recent study (but did extend the durability of anti-suicidal effects)^36^. A similar lack of effect on ketamine’ antidepressant durability has also been shown in 2 studies that used NMDAR antagonist riluzole after a ketamine infusion^37,38^. Broadly, as plasticity is attenuated by age, our results showing no effect of age on ketamine’s antidepressant durability are consistent with these reports showing no improvement of durability with administration of pharmacologic agents targeting the NMDAR and presumably NMDAR-dependent plasticity mechanisms.

Interestingly, a recent study examining the use of the mTORC1 inhibitor rapamycin following ketamine infusion did extend clinical durability effects, although the mechanistic implications of this phenomenon are unclear^39^. mTORC1 is a critical downstream signaling molecule of glutamatergic signaling via AMPA receptors resulting in synaptogenesis; several preclinical studies show that ketamine increases mTORC1 signaling^40,41^. The authors initially hypothesized that rapamycin would block ketamine’s antidepressant action (via blockade of mTORC1), but instead surprisingly reported both no effect of rapamycin administration on ketamine’s antidepressant efficacy and that rapamycin pretreatment actually lead to superior antidepressant durability. Thus, the existing literature as well as the current study suggest that plasticity mechanisms may be less important for maintaining ketamine’s antidepressant durability and that other mechanistic targets (e.g, inflammation, genetics, ketamine metabolites) may be of greater import.

Our study had several important limitations. We conducted a retrospective case-series analysis of veterans receiving clinical care at a VA ketamine clinic. As such, we are limited by the demographic and diagnostic heterogeneity of the sample, and lack the rigor of a randomized, controlled, prospective study design. Other demographic and treatment variables that may have covaried with age could have contributed to our reported effects. Additionally, it is possible that with more power, we would have demonstrated statistically significant age-dependent effects for infusions #5 and #6, which would have affected the interpretation of these results (namely showing that older patients had reduced total clinical response to the six-infusion ketamine series). Follow-up analyses with a larger controlled sample are needed to help to clarify this point. As BDI-II data were collected prior to each infusion (as per our standard clinical operations), we do not have BDI-II outcome data reflecting the effects of the sixth infusion in the series. While this does impact to an extent our reported group-level clinical outcomes for total response and durability effects, we do not expect the additional clinical improvement resultant from infusion #6 would manifestly change our reported correlations, particularly with respect to response durability at 3 weeks. Finally, our sample was highly treatment refractory (averaging ~9 failed antidepressant trials) which may affect the generalizability of these results to less treatment resistant populations.

Despite these limitations, this is the largest case-series to date examining the effects of patient age on antidepressant response trajectories and durability to serial ketamine infusions in patients with TRD. These results inform the current literature focusing on better characterizing the different phases of clinical response to serial ketamine infusions and possible mechanisms underlying ketamine’s antidepressant activity. This work contributes to efforts informing personalization and optimization of ketamine delivery parameters, as well as in developing mechanistic studies that can better capture the nuances of ketamine’s antidepressant action across a treatment course.

## Data Availability

Data available upon request of the corresponding author. This was not a clinical trial or prospective study so is not registered as a clinical trial. This was a retrospective case-series analysis.

